# A systematic review uncovers a wide-gap between COVID-19 in humans and animal models

**DOI:** 10.1101/2020.07.15.20147041

**Authors:** Salleh N. Ehaideb, Mashan L. Abdullah, Bisher Abuyassin, Abderrezak Bouchama

**Affiliations:** Experimental Medicine Department, King Abdullah International Medical Research Center/King Saud bin Abdulaziz University for Health Sciences, King Abdulaziz Medical City, Ministry of National Guard Health Affairs, Riyadh, Saudi Arabia

**Author notes:** **Correspondence** Dr. Abderrezak Bouchama, Experimental Medicine Department, King Abdullah International Medical Research Center/King Saud bin Abdulaziz University for Health Sciences, King Abdulaziz Medical City, Ministry of National Guard Health Affairs, Riyadh, Saudi Arabia.

## Abstract

**Background:** Animal models of COVID-19 have been rapidly reported after the start of the pandemic. We aimed to assess whether the newly created models reproduce the full spectrum of humans COVID-19.

**Methods:** We searched the Medline, as well as BioRxiv and MedRxiv preprint servers for original research published in English from January 1, to May 20, 2020. We used the search terms “COVID-19” OR “SARS-CoV-2” AND, “animal models”, “hamsters”, “nonhuman primates”, “macaques”, “rodent”, “mice”, “rats”, “ferrets”, “rabbits”, “cats”, and “dogs”. Inclusion criteria were the establishment of animal models of COVID-19 as an endpoint. Other inclusion criteria were assessment of prophylaxis, therapies, or vaccines, using animal models of COVID-19.

**Findings:** 13 peer-reviewed studies and 14 preprints met inclusion criteria. The animals used were nonhuman primates (n=13), mice (n=7), ferrets (n=4), hamsters (n=4), and cats (n=1). All animals supported high viral replication in the upper and lower respiratory tract associated with mild clinical manifestations, lung pathology and full recovery. Older animals displayed relatively more severe illness than the younger ones. No animal models developed hypoxemic respiratory failure, multiple organ dysfunction, culminating in death. All species elicited a specific IgG antibodies response to the spike proteins, which were protective against a second exposure. Transient systemic inflammation was observed occasionally in Rhesus macaques, hamsters, and mice. Notably, none of the animals unveiled cytokine storm or coagulopathy.

**Conclusions:** Most of the animal models of COVID-19 recapitulated mild pattern of human COVID-19 with full recovery phenotype. No severe illness associated with mortality was observed, suggesting a wide gap between COVID-19 in humans and animal models.

**Funding:** There was no funding source for this study.

## Introduction

Coronavirus disease 2019 (COVID-19) is a febrile respiratory illness due to a novel viral pathogen severe acute respiratory syndrome–coronavirus 2 (SARS-CoV-2)^1,2^. COVID-19 can progress to acute respiratory distress syndrome (ARDS), multiple organ dysfunction/failure (MOSD), culminating in death^2-4^.

SARS-CoV-2 is a beta coronavirus that binds with a high affinity to angiotensin-converting enzyme (ACE) 2 receptor and uses the transmembrane serine protease (TMPRSS) 2 as co-receptor to gain entry to cells^5-7^. ACE2 and TMPRSS2 are co-expressed in many tissues and organs, particularly the nasal epithelial cells and alveolar type II cells of the lungs, which may explain in part the easy transmission from person-to-person, and its dissemination within the body in severe and fatal cases^6-13^. Accordingly, SARS-CoV-2-induced COVID-19 has led to a pandemic that overwhelmed the capacity of most national health systems, resulting in a global health crisis^14^. So far, an estimated 11, 280 million persons in 188 countries were infected, of which 531.000 died^15^.

The clinical spectrum of COVID-19 is complex and has been categorized as mild, severe, and critical, representing 81, 14, and 5%^2,3^. The mild pattern comprises patients with either no signs and symptoms or fever and radiological evidence of pneumonia^3^. The severe pattern manifests as rapidly progressive hypoxemic pneumonia involving more than half of the lung with a full recovery phenotype^2,3^. The critical pattern consists of ARDS requiring respiratory assistance, MOSD that result in death in approximately half of the patients^2,3,16,17^. Mortality was associated with host factors such as old age, comorbidities, and immune response^4^.

Viral and immunopathological studies revealed distinct patterns between mild and severe or critical forms of COVID-19^4,17-25^. Both severe and critically ill patients displayed higher viral load in the upper respiratory tract than mild cases, together with delayed clearance overtime^17,18^. Likewise, they presented with lymphopenia due to a decrease in CD4+ and CD8+ T cells, as well as T cells exhaustion accompanied by a marked inflammatory response^20-25^. Pro- and anti-inflammatory cytokines and chemokines concentrations were increased systemically and locally in the lung and correlated with severity^20-22^. In contrast, in the mild illness, the lymphocyte count was normal, with no or minimal inflammatory response^19,22^. Together, these suggest that the viral load and dynamic together with the host inflammatory response may play a pathogenic role.

Clinical and post-mortem studies of fatal cases of COVID-19 demonstrated major alteration of coagulation and fibrinolysis^12,13^. This was associated with widespread thrombosis of small and large vessels, particularly of the pulmonary circulation contributing to death in a third of patients^26-32^. These observations suggest that dysregulated coagulation may be an important mechanism of COVID-19 morbidity and mortality^33^.

In this context, animal models appear crucial to a better understanding of the complex biology of COVID-19. Animal models of SARS-CoV-2-induced COVID-19 have been rapidly reported since the start of the pandemic^34^. However, whether they express the full phenotype of COVID-19, particularly the severe and critical patterns associated with lethality, remains to be determined. In this systematic review, we examined whether the newly created animal models reproduce the phenotype of human COVID-19. Moreover, we examined the knowledge generated by these models of COVID-19 including viral dynamic and transmission, pathogenesis, and testing of therapy and vaccines.

## Material and methods

### Search strategy and selection criteria

We conducted a systematic review according to the Preferred Reporting Items for Systematic Reviews and Meta-analysis (PRISMA) statement^35^ to identify studies describing the creation of an animal model of COVID-19 as an endpoint (Box1 and supplemental Box1).

### Results

The systematic search identified 101 studies and 326 preprints, of which 400 articles were excluded because they were reviews, non-original articles, or unrelated to the COVID-19 infection (Fig.1 and supplemental Table 1). 13 peer-reviewed studies and 14 preprints were included in the analysis.

**Figure 1.**
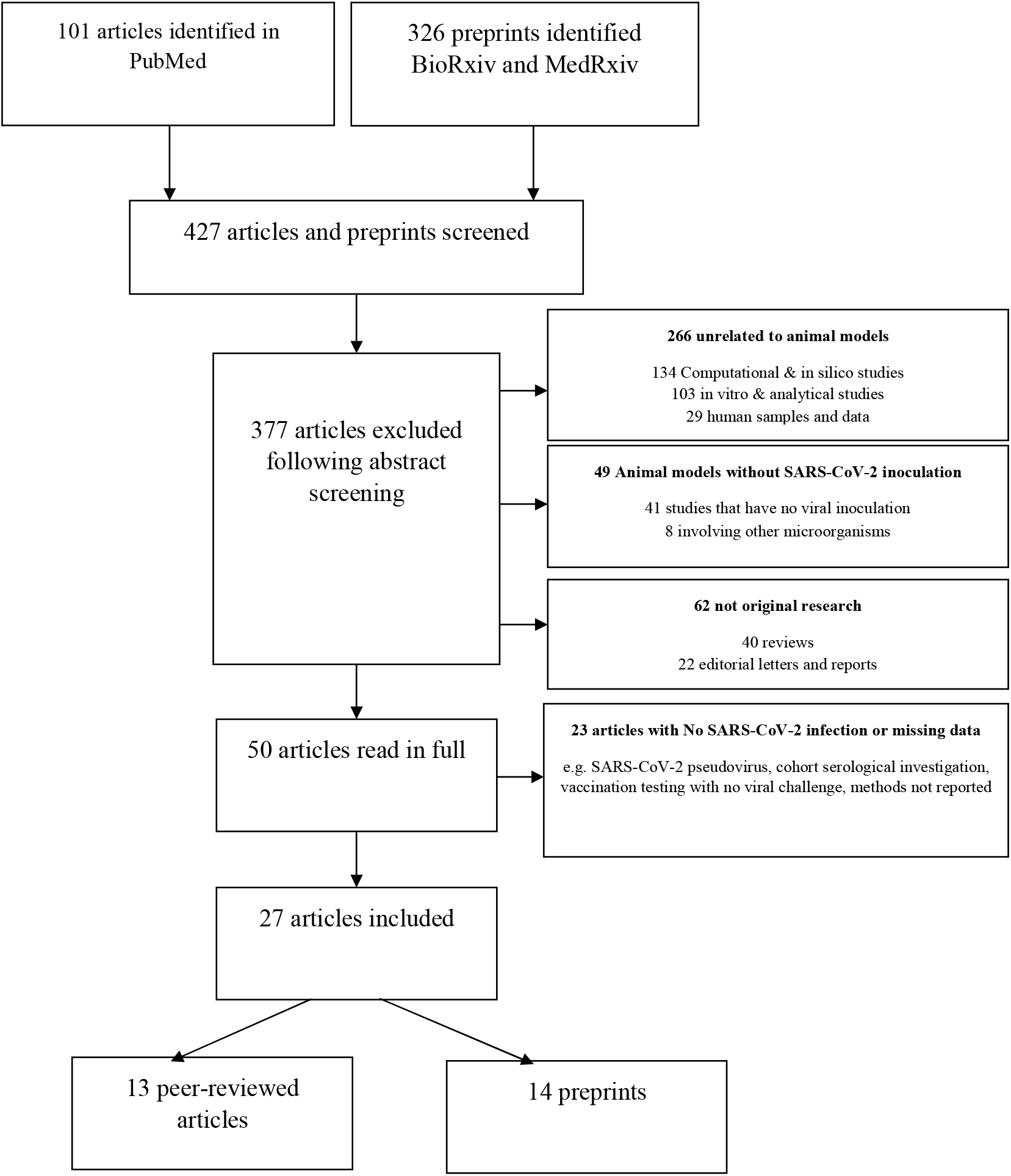
Flow diagram illustrating the process of studies selection *.

The studies used Old-world nonhuman primates (n=13)^36-48^, mice (n=7)^49-55^, hamsters (n=4)^55- 58^, ferrets (n=4)^59-62^, cats, and dogs (n=1)^62^ (Tables 1-4). Male and female, as well as young and old, were included but with no associated comorbidities. The aims were to investigate the pathogenesis of COVID-19 (n=15), testing drugs and vaccines (n=14), the host immune response (n=6), and the virus dynamic and transmission (n=4) (Tables 1-4).

**Table 1.**
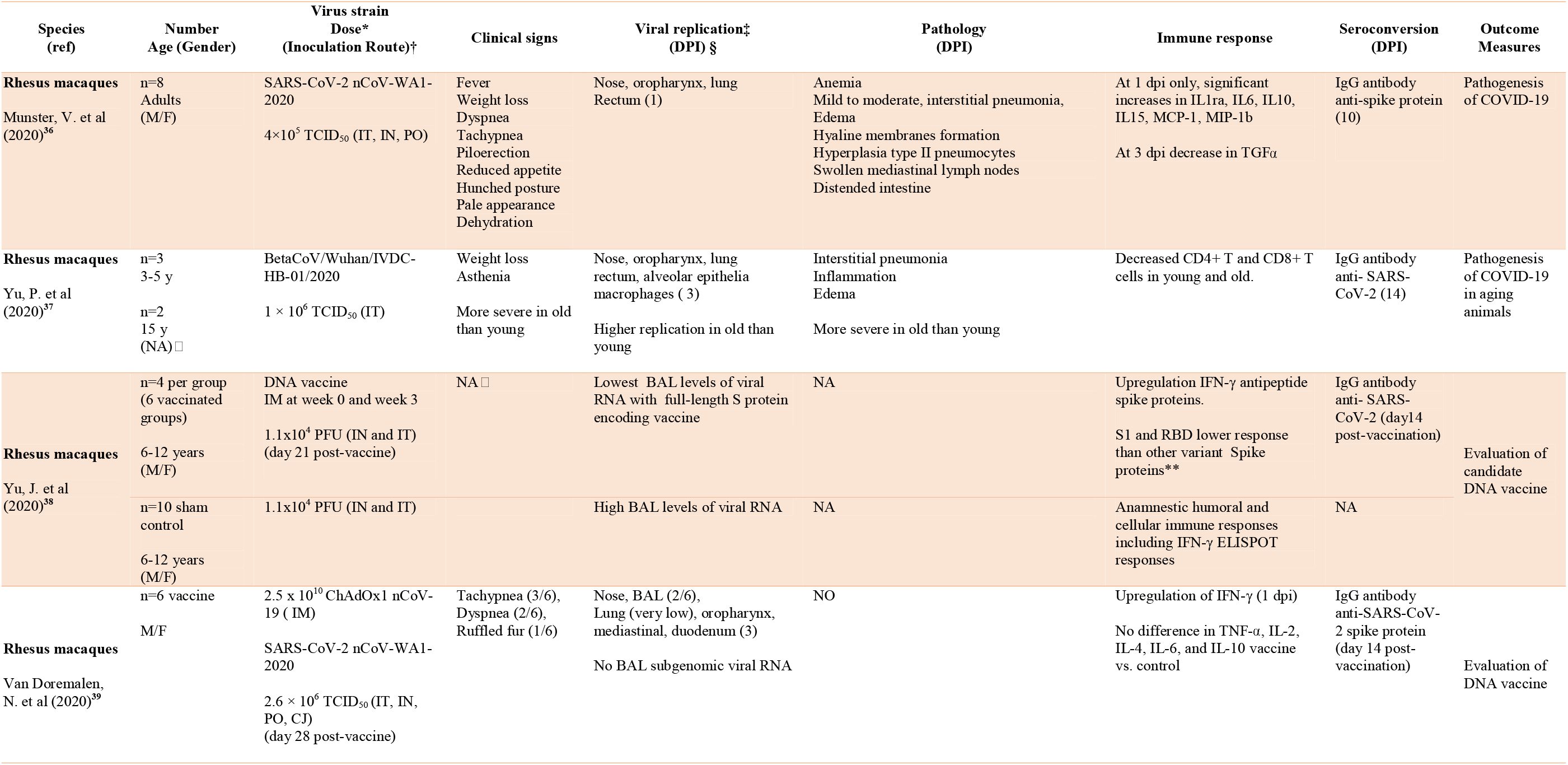
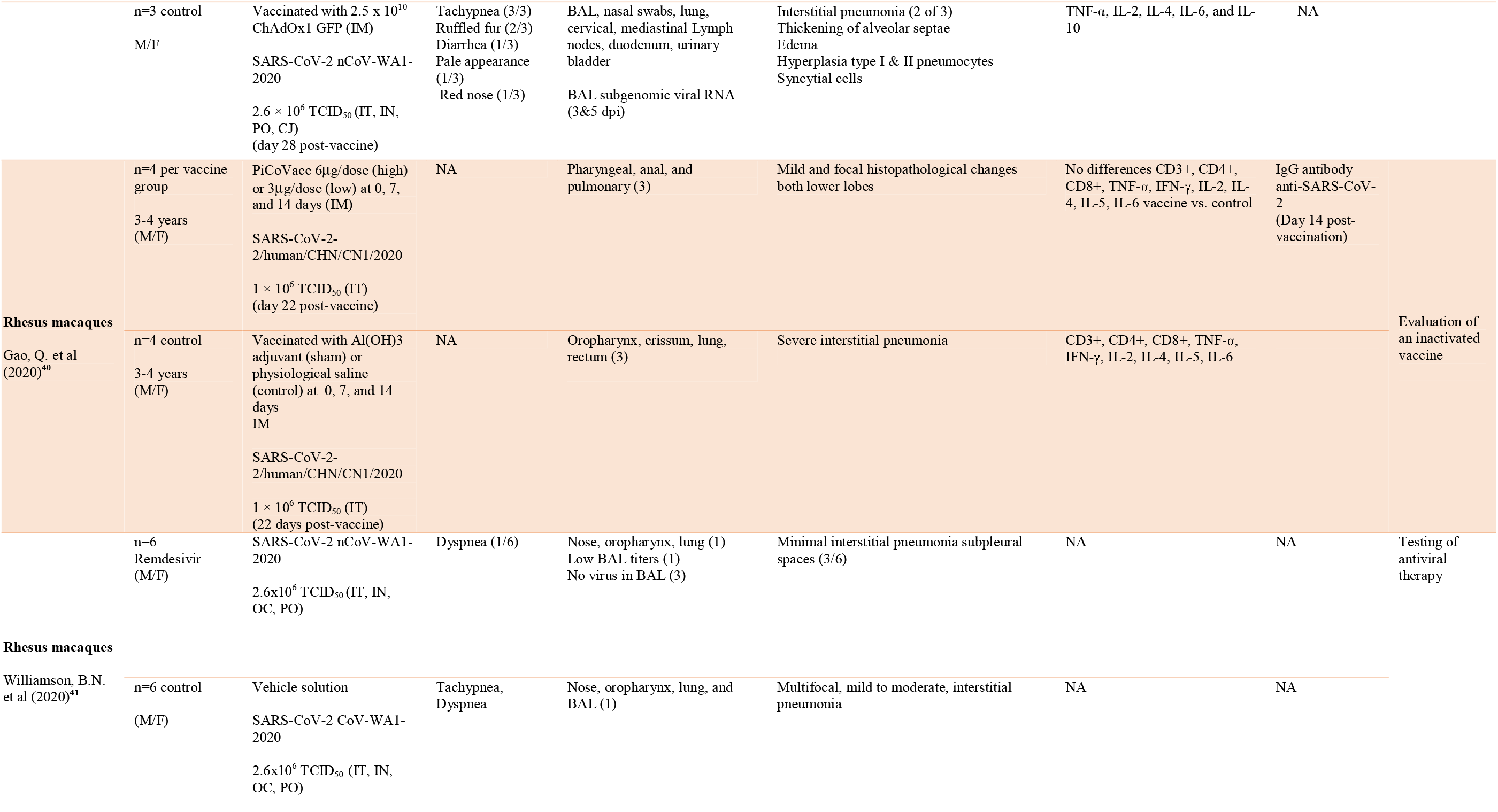
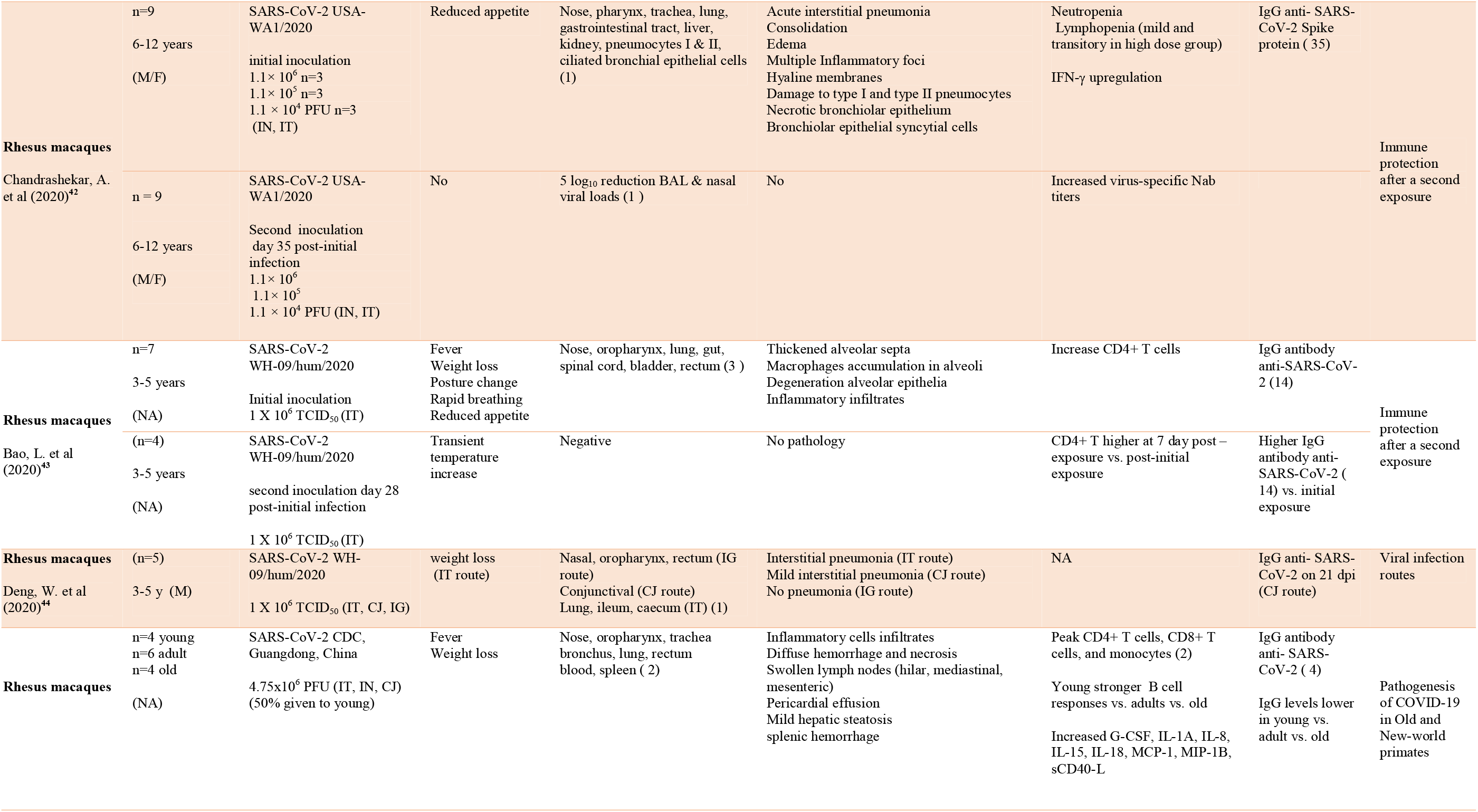
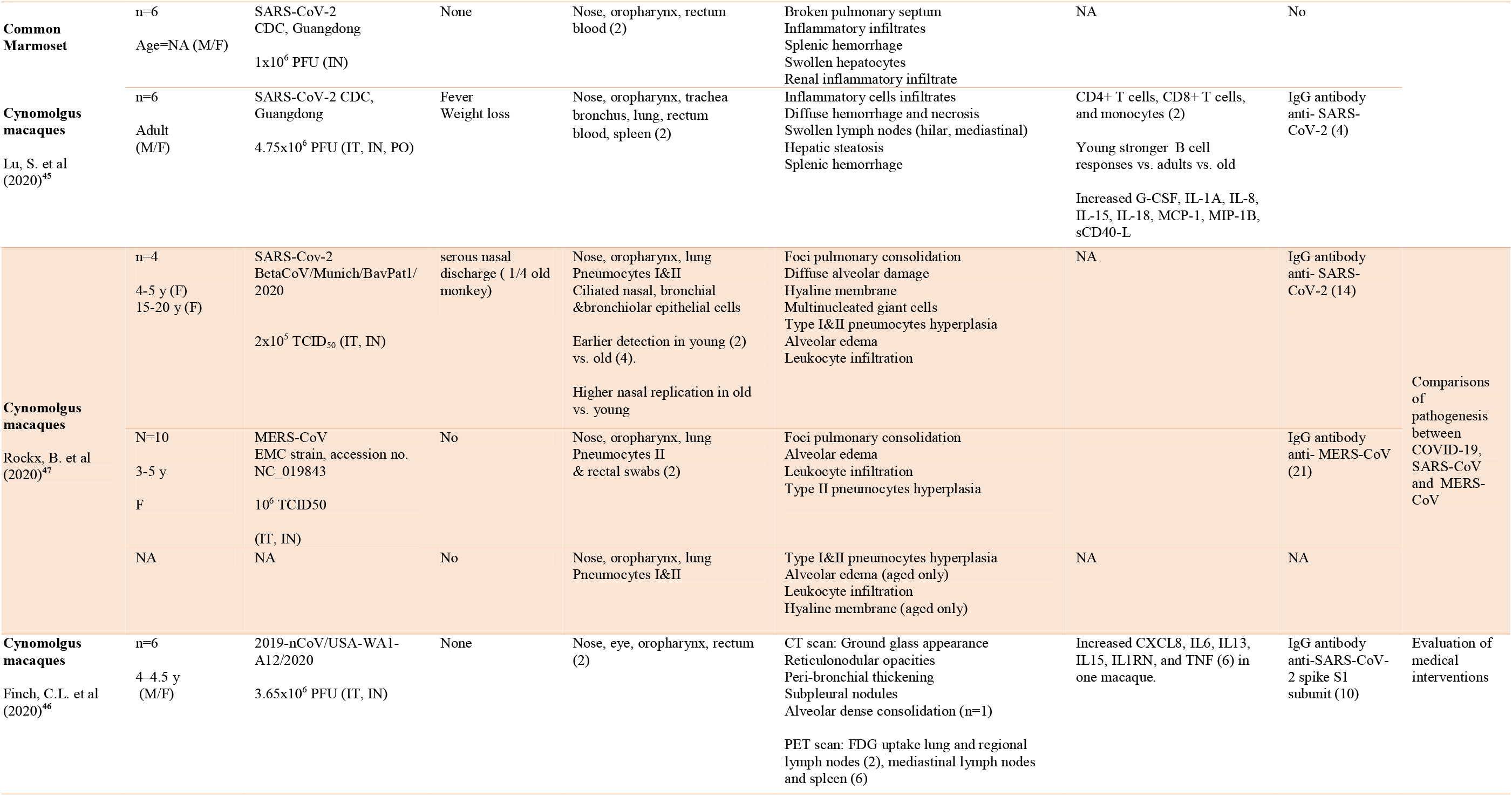
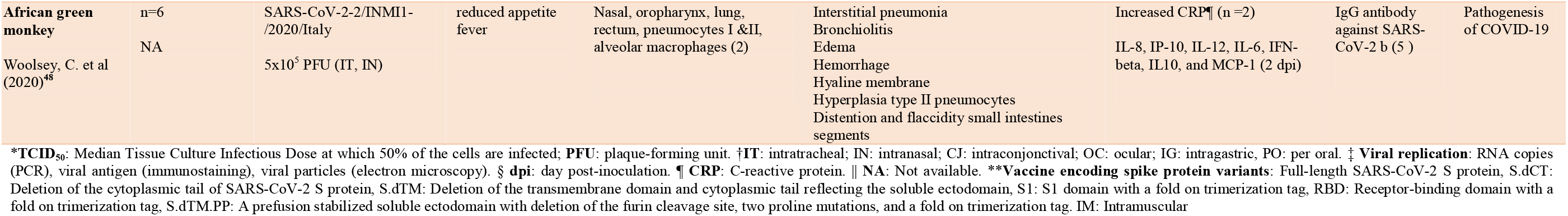
Summary of studies using nonhuman primate models of COVID-19

**Table 2.**
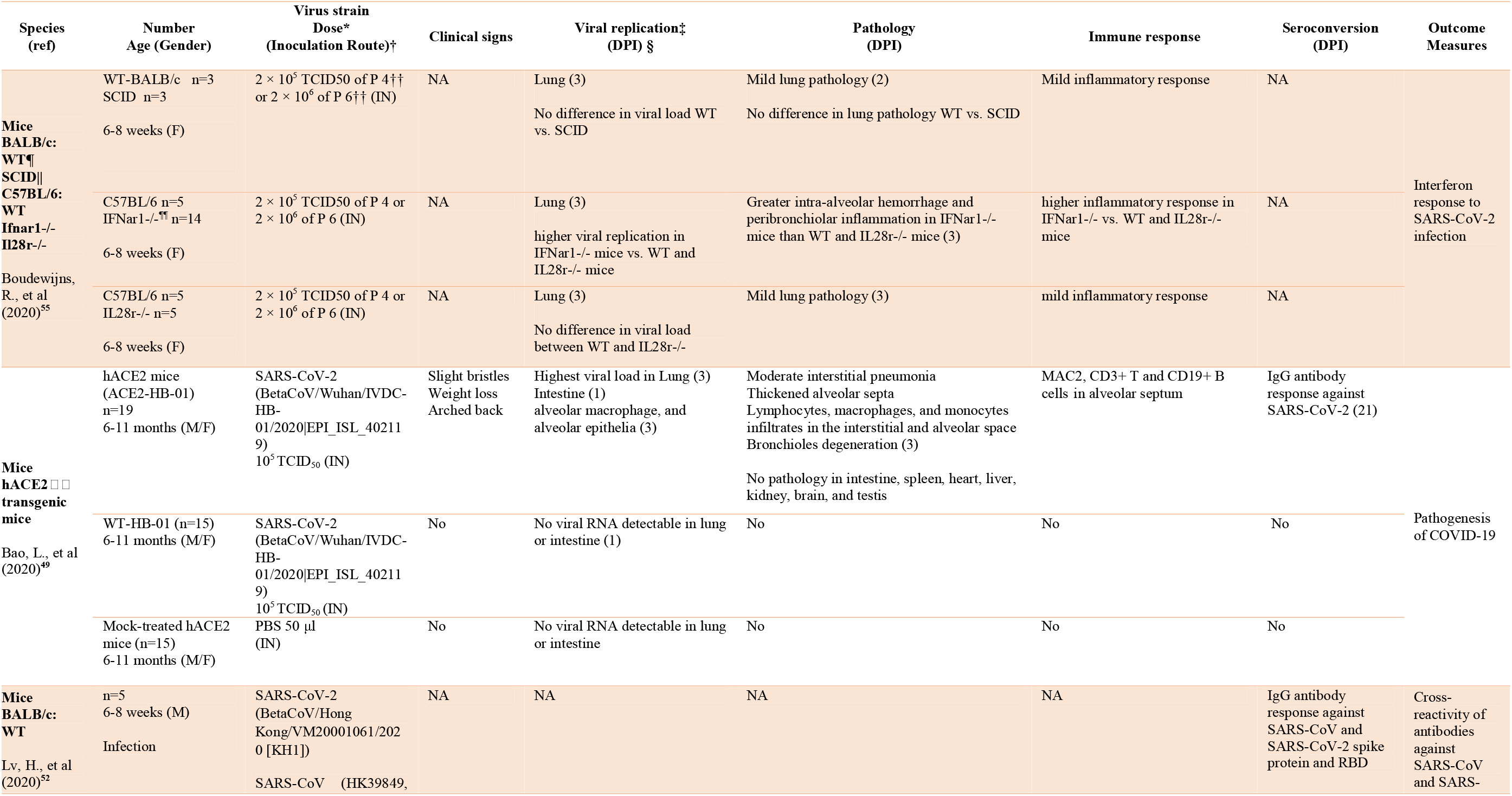
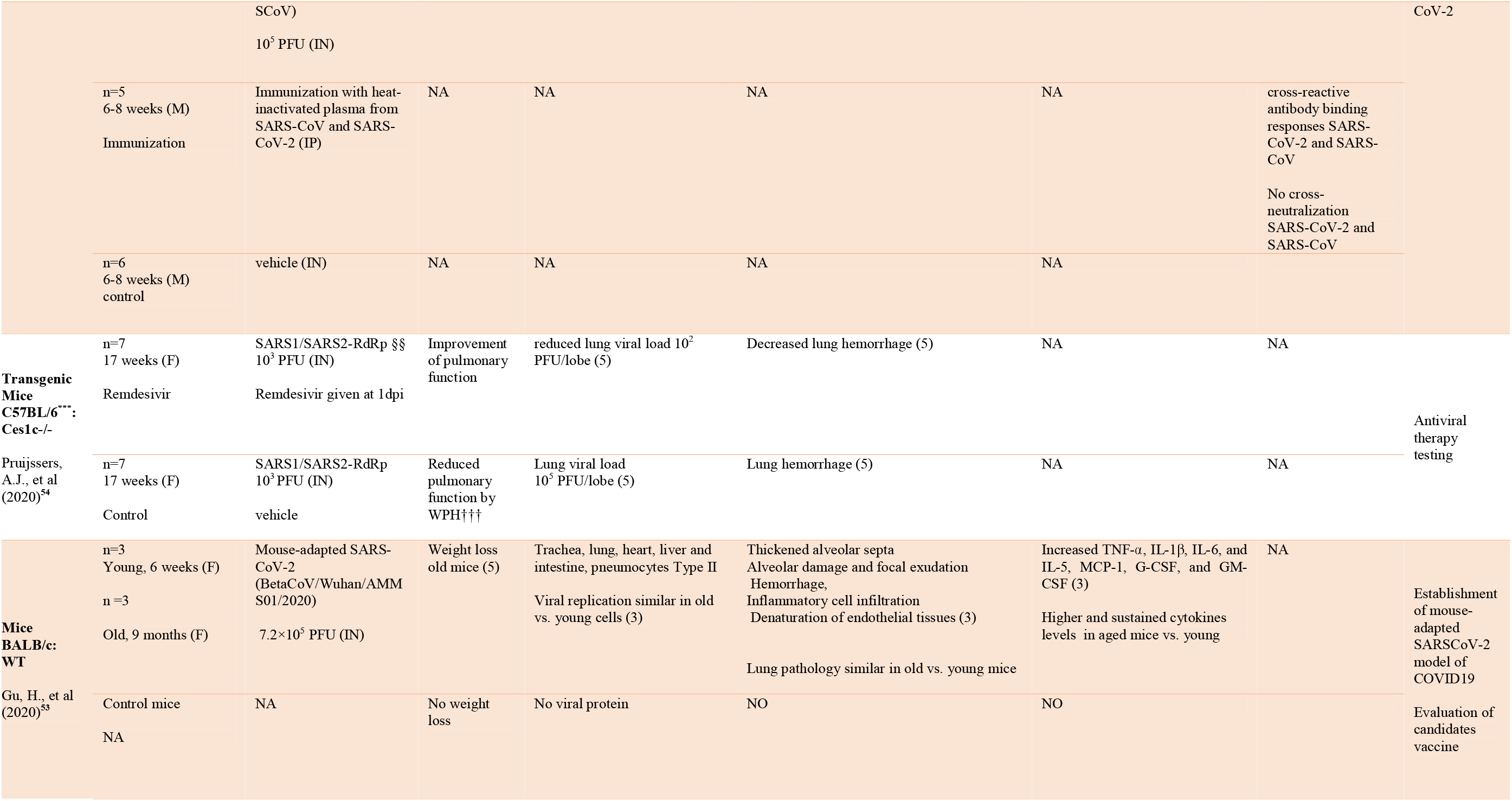
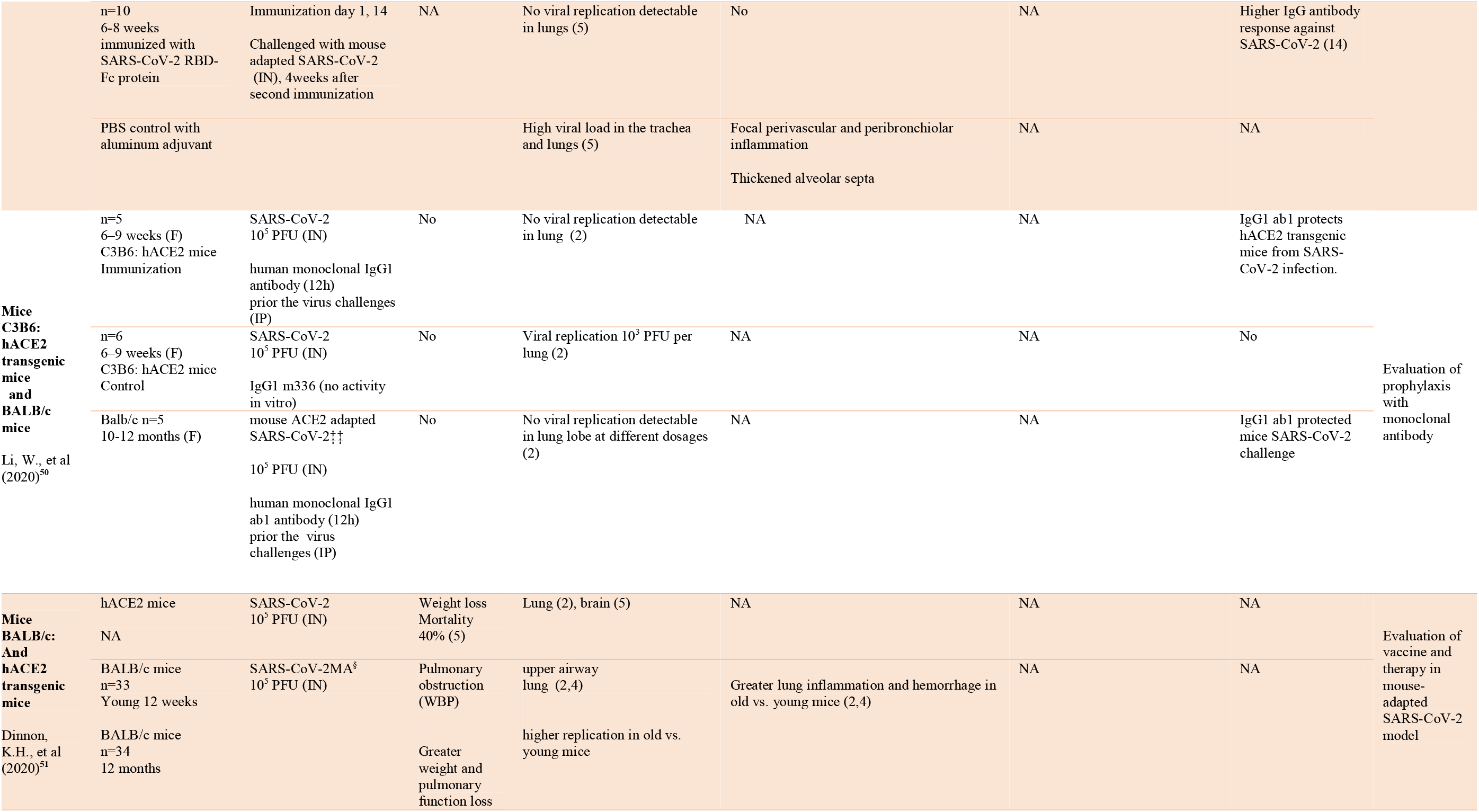
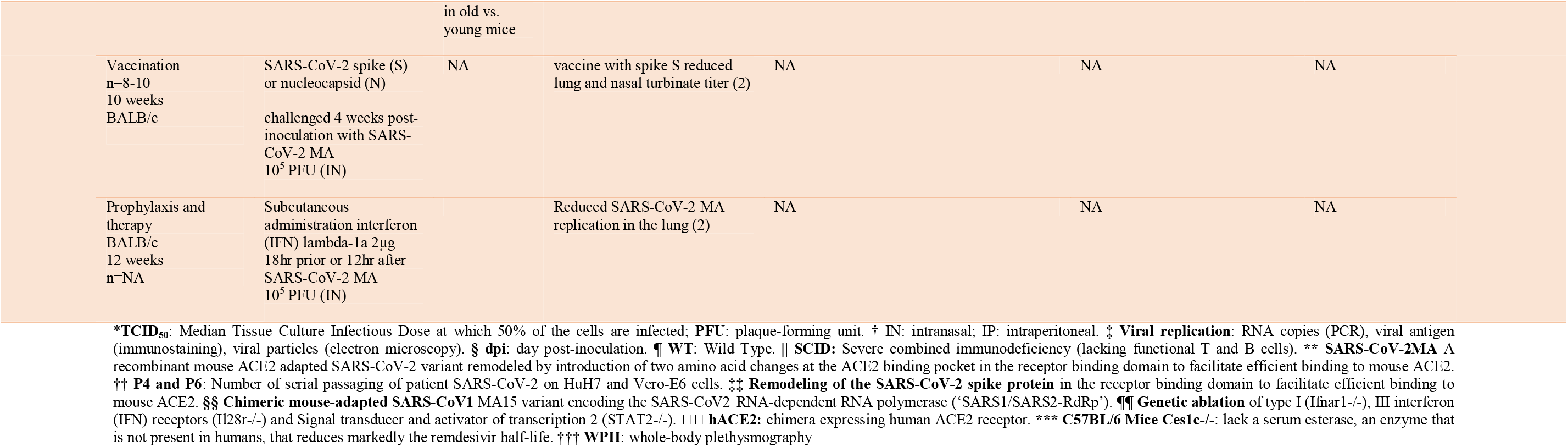
Summary of studies using mice models of COVID-19

**Table 3.**
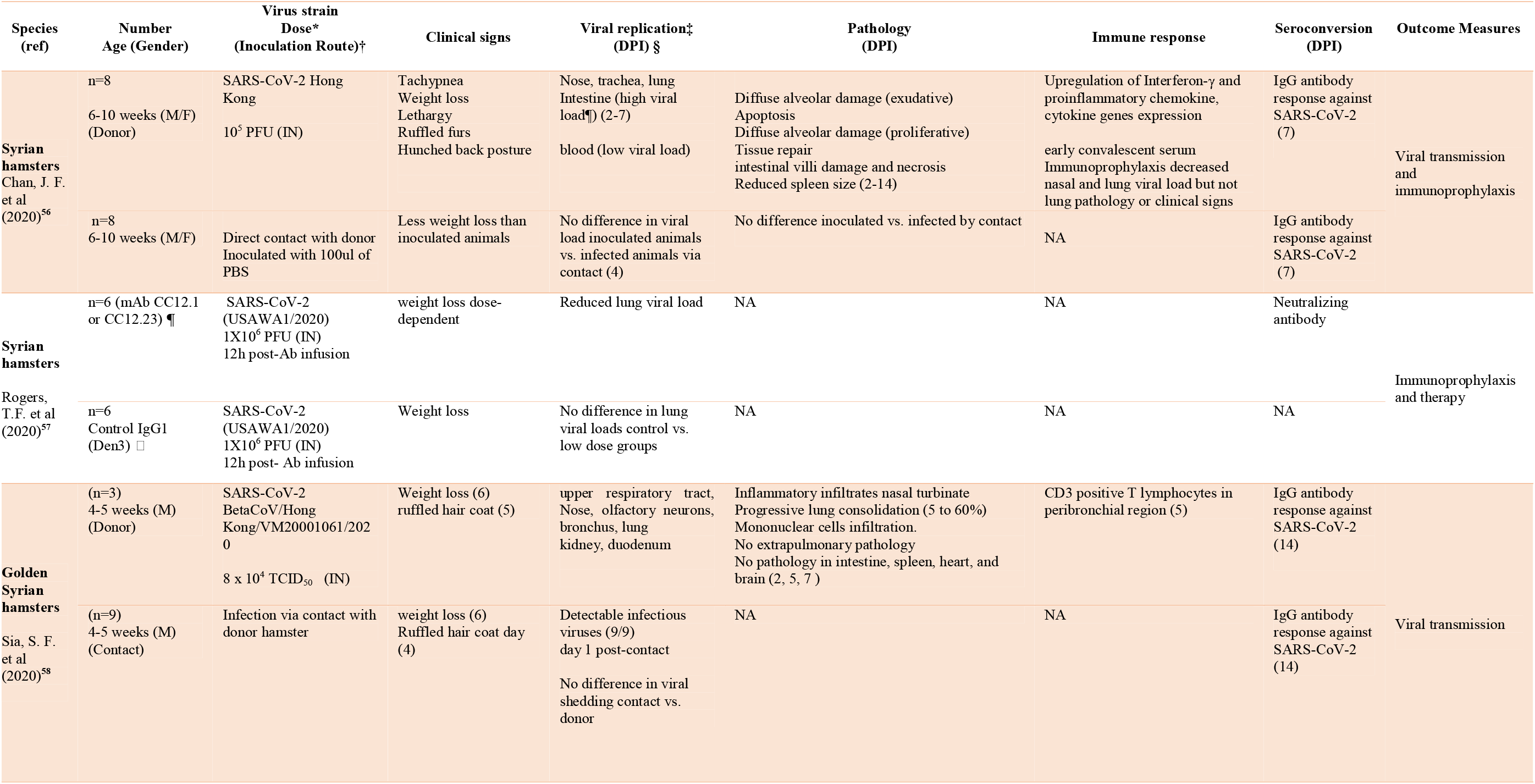
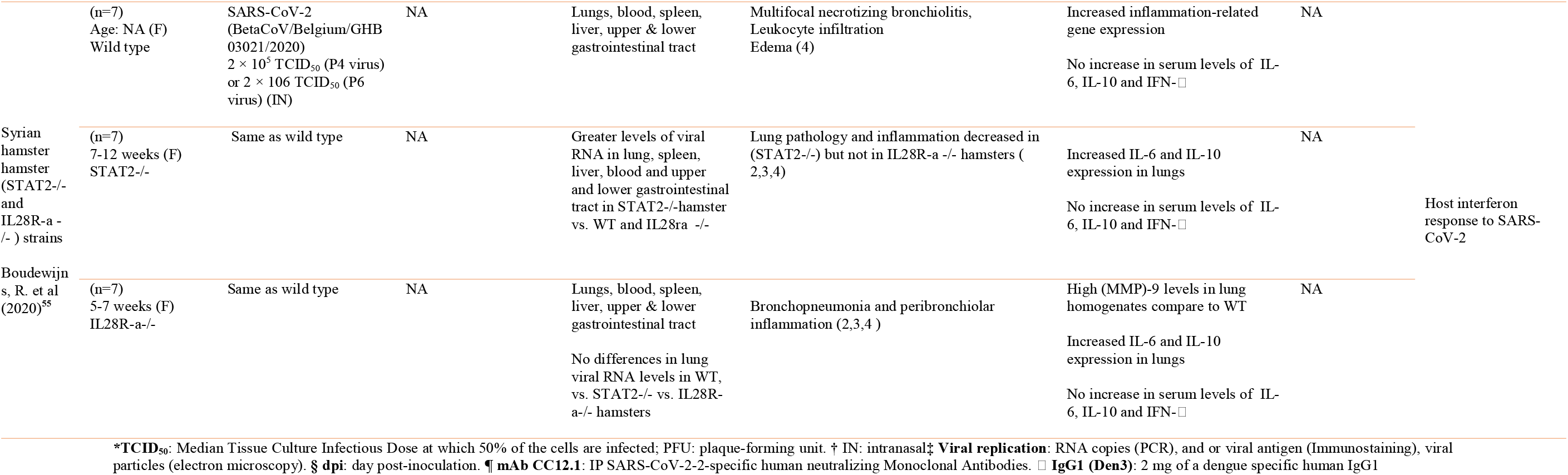
Summary of studies using hamsters models of COVID-19

**Table 4.**
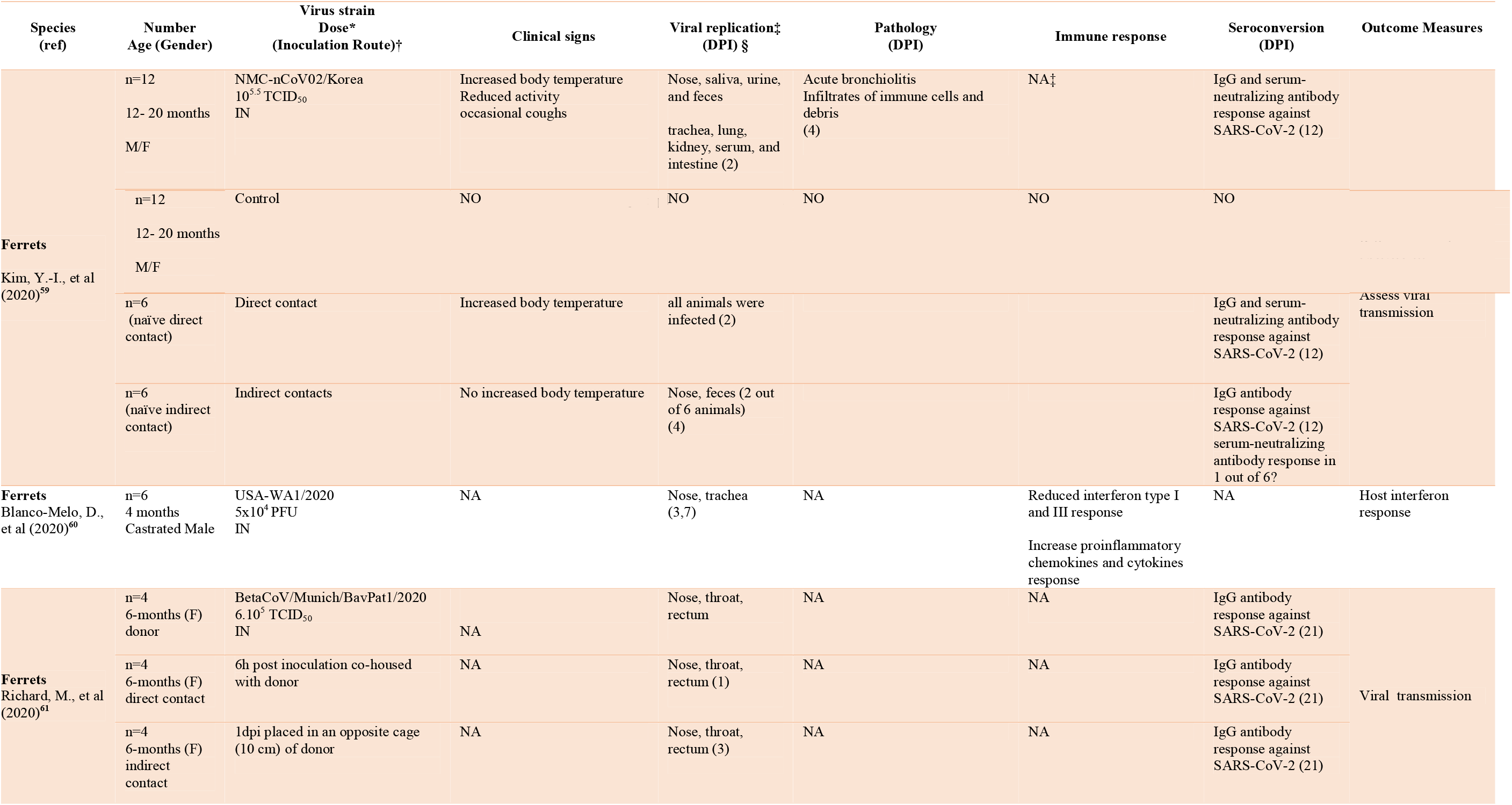
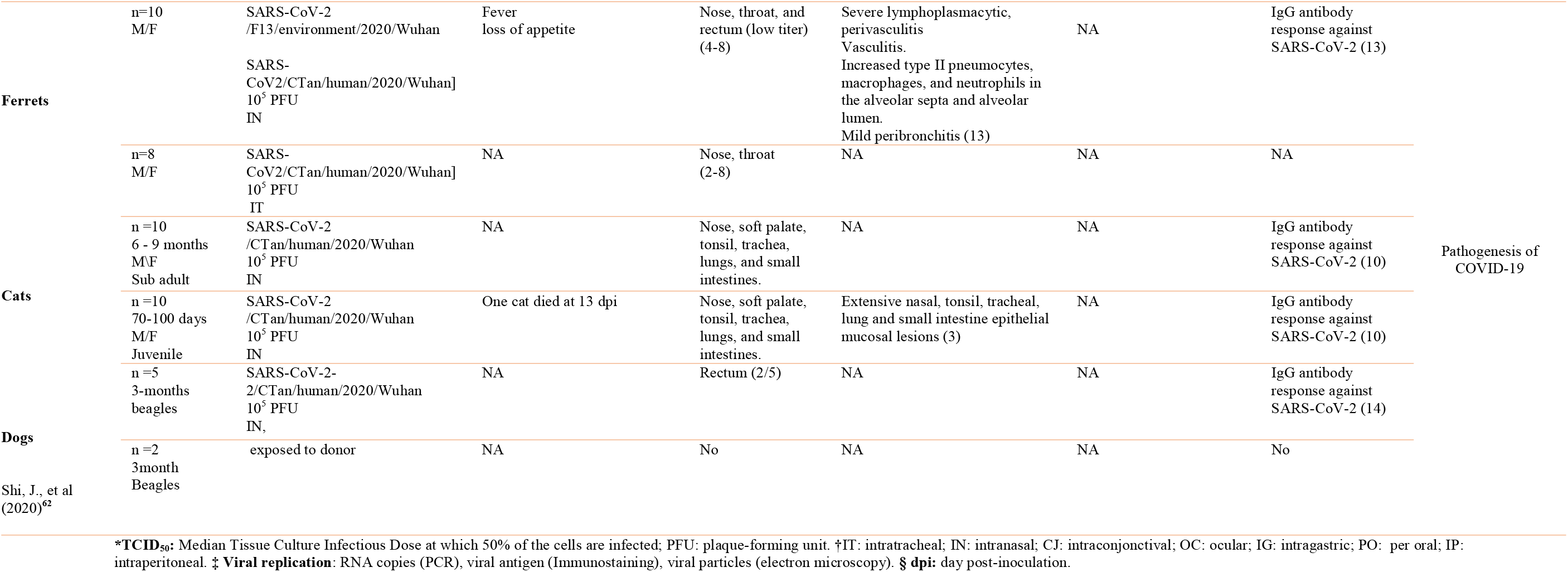
Summary of studies using ferrets, cats, and dog models of SARS-CoV-22 infection

Other animals studied included pigs, ducks, and chickens, however none of them supported SARS-CoV-2 replication, and hence will not be discussed further^62,63^.

All the experimental animals were inoculated with SARS-CoV-2 with various strains, doses, and route of administration that differed across studies (Table 1-4). Likewise, the time-point for tissue collection and pathological assessment were variables. These together precluded any comparisons between the animal models either intra-species or inter-species.

### Nonhuman primate models

#### Viral model

Rhesus macaques (n=10)^36-45^, Cynomolgus (n=3)^45-47^, and African Green model (n=1)^48^ from the Old World primates and common marmoset (n=1)^45^ from the New World were assessed as models for COVID-19 (Table 1). SARS-CoV-2 strains, dose and route of inoculation were different across studies. Infection through the intragastric, conjunctival and intratracheal routes elicited no lung pathology, mild and severe interstitial pneumonia, respectively, thus, emphasizing the relevance of the entry site of the virus in the expression of the disease^44^. The animals were euthanized at different time-points post-inoculation ranging from 3 to 33 days.

#### Phenotype

The animals displayed variable clinical manifestations from none to fever, altered respiratory patterns, and other general signs (Table 1). The clinical manifestations were not different between old and young macaques^45-47^. Structural and ultrastructural examination of the respiratory tract were also variables including mild to moderate interstitial pneumonitis, edema, foci of diffuse alveolar damage with occasional hyaline membrane formation and pneumocytes type II hyperplasia (Table 1). Old rhesus macaques exhibited more diffuse and severe interstitial pneumonia than young ones^46^. No extrapulmonary injury was documented except in two studies, which revealed inflammatory cells infiltrating the jejunum, and colon, steatosis of the liver, and alteration of myocardial fiber architecture with increased mitochondrial density^32,45^. No mortality was observed in any of the nonhuman primate models.

Comparisons between species of nonhuman primates was not possible except in one study, which suggested that rhesus macaques was superior to cynomolgus and common marmoset as models of human COVID-19^45^. Other comparisons suggested that SARS-CoV elicited more severe lung pathology than SARS-CoV-2 and MERS-CoV^47^ (Table 1).

#### Viral and host interaction

The virus replicated rapidly and at higher titers in upper airway and lung and intrathoracic lymph nodes in all four species. The virus was detected in pneumocytes type I and II, and ciliated epithelial cells of nasal, bronchial and bronchiolar mucosa. This differs from MERS-CoV where the virus was mainly present in type II pneumocytes^45^ (Table1). Replication of the virus was also demonstrated in jejunum, colon, and rectum^27,36,48^. Viral genome was detected in the blood of rhesus macaques, cynomolgus, and marmoset in one study^45^. Viral replication of nasopharyngeal as well as anal swabs, and lung in old macaques was higher than in young ones^46,47^.

SARS-CoV-2 infection-induced IgG antibodies response against the SARS-CoV-2 spike was noted in all species^36,45,47,48^ except in marmoset^45^. The antibodies were protective against a second exposure to the virus. There was no difference between males and females; however, young rhesus macaques had lower antibodies titers than the old macaques^45^. The innate immune response to SARS-CoV-2 infection was variable with normal, high or low leucocytes and lymphocytes counts^36,45^. Occasional reduction of CD4^+^ and CD8^+^ T cells concentrations was documented^36^ as well as transitory release of various cytokines, and chemokines at different days post-inoculation^36,45,48^.

#### Drugs and vaccines

DNA and inactivated virus-based vaccines were evaluated and showed protection in these nonhuman primates. However, the DNA vaccine did not reduce the virus presence in the upper airway, while there was a residual small interstitial pneumonitis in the macaques that received inactivated virus^39,40^. This suggests that none of the vaccines tested so far, displayed a comprehensive protection against SARS-CoV-2 infection. Several candidate DNA vaccines based in various forms of the SARS-CoV-2 Spike (S) protein were also tested in rhesus macaques^38^. The findings revealed that only the vaccine encoding the full-length (S) offered optimal protection against SARS-CoV-2^64^. Nonhuman primates served also for the evaluation of antiviral therapies and medical interventions such as CT- and PET-scanners^46^.

### Mouse models

#### Viral model

Wild type mice (BALB/c, C57BL/6); immunodeficient mice (SCID); chimeric mouse expressing human angiotensin-converting enzyme 2 (hACE2), and the RNA-dependent RNA polymerase (SARS1/SARS2-RdRp) were evaluated as models of COVID-19 (Table 2). Moreover, knockout (KO) mice were generated to test specific immunological pathways or therapy, including ablation of type I (IFNar1-/-) and III interferon (IFN) receptors (IL28r-/-), signal transducer and activator of transcription 2 (STAT2-/-); and serum esterase (Ces1c-/-).

Patients’ isolates of SARS-CoV-2 from different sources and variable times of passaging on various cell cultures or BALB/c mice were employed (Table 2). The multiple passaging on whole animal adapted the virus to the murine ACE-2^53^. SARS-CoV-2 was also modified through the remodeling of the spike receptor-binding domain (RBD), which enhanced its binding efficiency to murine ACE2^51^.

#### Phenotype

The clinical signs and symptoms varied from none to mild weight loss, arched back, and slight bristles. Whole-body plethysmography was used to measure the respiratory function of the animals and showed a mild to moderate reduction in old more than in young (Table 2). Likewise, the pathological changes varied according to the experimental models and included peribronchiolar inflammation, lung edema, moderate multifocal interstitial pneumonia, lymphocytes infiltration, and intra-alveolar hemorrhage. Survival of hACE2 mice was decreased at 5 day post-inoculation and was attributed to high viral replication in the brain, while it was minimal in the lung, suggesting a different pathogenic mechanism of death from human COVID-19^51^. Wild type mice showed no pathology as compared to hACE2 mice, indicating that lack of human ACE2 receptor cannot be infected or inefficiently with SARS-CoV-2^49,55^. On the other hand, mouse-adapted SARS-CoV-2 exhibited more severe pathology, particularly in the aged mouse than hACE2 transgenic mouse, suggesting that these models may be more relevant for the study of human COVID-19^51,53^. However, whether the pathogenesis induced by the mouse-adapted SARS-CoV-2 is translatable to human warrants further studies^51,53^.

#### Viral and host interaction

The virus replicated to high titers in the upper and lower respiratory tract in most of the genetically modified mice models but not in wild type. Viral replication was detected outside the respiratory tract in the intestine of hACE2 mice^49^ as well as in the liver, and heart in mouse modified SARS-CoV-2 RBD^51^. Increased viral replication in KO mice IFNar1-/- suggested that interferon limited the viral replication^55^.

Specific IgG antibodies against SARS-CoV-2 were documented in two studies (Table 2). The IgG antibodies were found to cross-react in their binding to the spike protein of SARS-CoV, however, with no cross-neutralization, hence suggesting the conservation of the same spike protein epitopes among coronaviruses^52^. Proinflammatory cytokines and chemokines were demonstrated in mouse-adapted SARS-CoV-2, and KO mouse (Table 2). The inflammatory response was significantly higher in the old than young mice.

#### Drugs and vaccines

Antiviral therapies, including remdesivir^54^, IFN lambda^51^, and human monoclonal IgG1 antibody against RBD^49^, were tested in these mouse models and produced a protective effect. Likewise, vaccines using viral particles expressing SARS-2-S protein^51^, or an RBD-based vaccine were tested and showed protection^54^.

### Hamsters

#### Viral model

Wild type Syrian hamsters and knockout hamsters for signal transducer and activator of transcription 2 (STAT2-/- lacking type I and III interferon signaling) and interleukin 28 receptors (IL28r-/- lacking IFN type III signaling) were reported as models for COVID-19. Patients isolate of SARS-CoV-2 from different sources and different passages on various cell cultures were used (Table 3). SARS-CoV-2 was administered intranasally at different titers to anesthetized hamsters. Viral transmission between hamsters was demonstrated either through direct contact or indirectly via airborne transmission.

#### Phenotype

The clinical manifestations included weight loss, which was consistently observed. Other clinical signs and symptoms such as rapid breathing, lethargy, ruffled furs, and hunched back posture were reported in one study^56^. The histopathological findings were variables according to the experimental models and ranged from lung consolidation to multifocal necrotizing bronchiolitis, leukocyte infiltration, and edema. STAT2 -/- hamsters exhibited attenuated lung pathology as compared with IL28R-a -/- hamsters suggesting that interferon type I response contributed to the injury^55^.

#### Viral and host interaction

The virus replicated to high titer in the upper and lower respiratory tract in most of the hamsters’ models. Viral replication was detected in blood and kidney with low concentration (Table 3). STAT2-/- hamsters had higher titers of infectious virus in the lung, viremia, and high levels of viral RNA in the spleen, liver, and upper and lower gastrointestinal tract in comparison with wild type and IL28R-a -/- hamsters. This suggests that IFN type I response limited the replication of SARS-CoV-2 infection. The mechanisms of this conflicting effect namely restricting the virus replication, while in the same time contributing to lung injury remains unclear. Specific IgG antibodies against SARS-CoV-2 were documented in the sera of hamsters at different time-points from virus inoculation ranging from 7 to 21 days. Increased expression of proinflammatory and chemokine genes was demonstrated in the lungs of the SARS-CoV-2 infected animals, however with no increase in circulating levels of proteins such as TNF, interferon-γ, and IL-6.

#### Drugs and vaccines

Immunoprophylaxis with early convalescent serum achieved a significant decrease in viral lung load but not in lung pathology^56^.

### Ferrets, cat, and dog

#### Viral model and phenotype

Ferrets, cats, and dogs were administered intranasally or intratracheally with various doses and strains of SARS-CoV-2 (Table 4). Ferrets displayed elevated body temperature for several days associated with signs that differed according to the studies. These include decreased activity and appetite, sporadic cough, and no body weight loss^59-62^. No clinical signs were reported either in cats or in dogs.

#### Histopathological changes

Ferrets exhibited acute bronchiolitis^60,62^, with perivasculitis and vasculitis^62^, but with no discernible pneumonia. Cats disclosed lesions in epithelial nasal, tracheal, and lung mucosa (Table 2).

#### Viral and host interaction

The virus replication and shedding were demonstrated in the upper airways and rectal swabs in ferrets and cats, but the extent to other tissues varied in ferrets from none to multiple organs, including lung, blood, and urine. No viral RNA was detected in cats’ lungs. Dogs showed RNA positive rectal swab but none in upper or lower airways. Viral transmission between ferrets and cats was demonstrated either through direct contact^54^ or indirectly via airborne route^61^.

Ferrets, cats, and dogs exhibited specific antibody response against SARS-CoV-2^59,61,62^. A study of the ferret immune response to SARS-CoV-2 revealed a subdued low interferon type I and type III response that contrasts with increased chemokines and proinflammatory cytokine IL6, which is reminiscent of the human response^60^.

## Discussion

This systematic review of experimental animal models of SARS-CoV-2 induced-COVID-19 identified 13 peer-reviewed studies and 14 preprints that reported data on nonhuman primates^36-48^, mice^49-55^, hamsters^55-58^, ferrets^59-62^, cats and dogs^62^ models of COVID-19. The main findings indicate that most of the animal models could mimic many features of mild human COVID-19 with a full recovery phenotype^3^. They also revealed that older animals display relatively more severe illness than the younger ones^37,45,47,51,53^, which evokes human COVID-19^3,65^. However, none of the animal models replicated the severe or critical patterns associated with mortality as observed in humans with COVID-19^3^.

The results of this systematic review are consistent with studies of animal models of SARS-CoV-2 and MERS-CoV, which failed to replicate the full spectrum of humans’ illness^66,67^. Nonetheless, several features of mild COVID-19 in humans could be mirrored. High viral titers in the upper and lower respiratory tract and lung pathology were demonstrated in both large and small animal models. The pathology encompassed mild interstitial pneumonia, consolidation, and diffuse alveolar damage, albeit localized to small lung area, edema, hyaline membrane formation, and inflammation. SARS-CoV-2 elicited specific antibodies response against various viral proteins in the sera of most of the animal models.

This systematic review revealed that none of these newly established animal models replicated the common complications of human COVID-19 such as ARDS and coagulopathy^26-32,65,68,69^. ARDS can be particularly severe and results in refractory hypoxemia requiring maximum respiratory supportive measures in intensive care unit^65,68,69^. The coagulopathy can lead to severe complications such as massive pulmonary embolism, cerebrovascular stroke, and mesenteric infarction, including in younger people^26,27,31,32^. The pathology underlying these two complications were recently revealed by post-mortem studies disclosing diffuse alveolar damage involving the whole lung, hyaline membrane formation, and infiltration with inflammatory cells, thus leaving no air space open for ventilation^12,13,64,70,71^. It also detected the presence of diffuse and widespread thrombosis in the micro- and macro-circulation, including the pulmonary circulation compromising the lung perfusion^12,13^. This double hit affecting the ventilation and perfusion simultaneously underlies the intractable hypoxemia that contributed to the high mortality. None of the animal models replicated the respiratory failure, thromboembolic manifestations, and their pathological expression, hence, indicating that a wide gap separates the animal models from the full spectrum of COVID-19 in humans.

The mechanisms of the lung injury and coagulopathy are not well understood, although several known pathways were postulated including cytokines storm leading to upregulation of tissue factor^20-22^, activation/injury of the endothelium infected by the virus^29,68,72^, complement activation^73^, alveolar hypoxia promoting thrombosis^74^, and autoantibodies against phospholipid and lupus anticoagulant^75,76^ modulating the hemostasis and coagulation cascade directly. Hence, the development of animal models that replicate the dysregulation of the inflammation and coagulation could be important, as these would allow the deciphering of the intimate mechanisms at play. This, in turn, may aid in identifying therapeutic targets and the testing of immunotherapy, anticoagulation, and thrombolytic interventions and thereby may improve the outcome.

Both antiviral and vaccine therapies were tested in rhesus macaques and mice infected with SARS-CoV-2^39-41^. The antiviral drug stopped the viral replication and improved the pneumonitis^41,54^. The vaccines induced an increase in titers of neutralizing antibodies in the sera that correlated with the decrease of viral replication and prevented the lung pathology^38-40^. These results represent a substantial proof of the concept of antiviral or vaccine efficacy against SARS-CoV-2 in animal models. However, because of the lack of overt clinical illness, the rapid clearance of the virus, and spontaneous improvement of the pneumonitis without lethality, the models do not permit the full assessment of the duration of the protection of the vaccines, or the effect of antiviral therapy on survival.

Since the emergence of SARS-CoV infection in 2003^77^, followed by the MERS-CoV in 2012^78^, and now with COVID-19, researchers have not been able to develop a model of coronavirus infection that reproduces the severity and lethality seen in humans^66,67^. One of the well-known reasons lies in the difference of ACE-2 receptor binding domain structure across species^79^. Human and old-world primates have conserved a comparable structure that allows binding with high affinity to the SARS-CoV-2^79^. The new-world primates, hamsters, ferrets, and cats maintained an intermediate affinity, while mice exhibit very low affinity^79^. The latter explains why wild-type mouse does not support SARS-CoV-2 replication, and hence, the necessity to create a chimera that expresses human ACE-2, to enable the use of this species as model of COVID-19^49^. More recently, a study applying single-cell RNA sequencing to nonhuman primate uncovered another explanation that may underlies the difference between old-world primates and humans in expressing the complex phenotype of COVID-19^80^. The study reveals that the cellular expression and distribution of ACE2 and TMPRSS2 which are essential for virus entry in the cells and its spread inside the body differ in the lung, liver, and kidney between the two species. ACE2 expression was found lower in pneumocytes type II and higher in ciliated cells in nonhuman primate lung as compared to humans^39^. This is particularly significant as type II pneumocytes are critical targets of SARS-CoV-2 in humans and the pathogenesis of lung injury/damage. Finally, the innate immune response including the defense system against viruses diverged during evolution both at the transcriptional levels and cellular levels, which may also explain why the SARS-CoV-2 hardly progresses in these animals outside the respiratory system^81^. Taken together, these fundamental differences represent a real challenge to the successful development of an animal model that reproduces human COVID-19.

This systematic review has few limitations. First, it is the high number of preprints included in this study that have not been peer-reviewed. Second, the animal models from the same species were difficult to compare across studies, as they used different viral strain, inoculum size, route of administration, and timing of tissue collection.

In conclusion, this systematic review revealed that animal models of COVID19 mimic mild human COVID-19, but not the severe form COVID-19 associated with mortality. It also disclosed the knowledge generated by these models of COVID-19 including viral dynamic and transmission, pathogenesis, and testing of therapy and vaccines. Likewise, the study underlines the distinct advantages and limitations of each model, which should be considered when designing studies, interpreting pathogenic mechanisms, or extrapolating therapy or vaccines results to humans. Finally, harmonization of animal research protocols to generate results that are consistent, reproducible, and comparable across studies is needed.

## Data Availability

All supplemental data are included.

## Box 1.

### Search strategy and selection criteria

We searched the Medline, as well as BioRxiv and MedRxiv preprint servers for original research describing or using an animal model of SARS-CoV-2 induced COVID published in English from January 1, 2020, to May 20, 2020. We used the search terms “COVID-19” OR “SARS-CoV-2” AND, “animal models”, “hamsters”, “nonhuman primates”, “macaques”, “rodent”, “mice”, “rats”, “ferrets”, “rabbits”, “cats”, and “dogs”. The preprint servers were included in the search as the field of COVID-19 is developing quickly. Inclusion criteria were establishment of animal models of COVID-19 as endpoint. Other inclusion criteria were assessment of prophylaxis, therapies, or vaccines, using animal models of COVID-19. 101 studies and 326 preprints were screened, of which 13 peer-reviewed studies and 14 preprints were included in the final analysis (Fig.1). The variables extracted were the population type, study aim, the virus strain used, clinical response, pathology, viral replication, and host response as well as the effects of prophylaxis, drugs, or vaccines. The outcomes were organized according to species and categorized into phenotype (signs or symptoms; histopathology, time-course of the illness and outcome), viral (titer in each tissue organ, detection methods, duration of positivity), and host response (dynamic of seroconversion, inflammatory and hemostatic markers), therapy and vaccine (efficacy, and safety).

## Reference

1. Zhou P, Yang XL, Wang XG, et al. A pneumonia outbreak associated with a new coronavirus of probable bat origin. Nature 2020; 579(7798): 270–3.

2. Zhu N, Zhang D, Wang W, et al. A Novel Coronavirus from Patients with Pneumonia in China, 2019. The New England journal of medicine 2020; 382(8): 727–33.

3. Wu Z, McGoogan JM. Characteristics of and Important Lessons From the Coronavirus Disease 2019 (COVID-19) Outbreak in China: Summary of a Report of 72L314 Cases From the Chinese Center for Disease Control and Prevention. Jama 2020.

4. Zhang X, Tan Y, Ling Y, et al. Viral and host factors related to the clinical outcome of COVID-19. Nature 2020.

5. Wrapp D, Wang N, Corbett KS, et al. Cryo-EM structure of the 2019-nCoV spike in the prefusion conformation. Science (New York, NY) 2020; 367(6483): 1260–3.

6. Hoffmann M, Kleine-Weber H, Schroeder S, et al. SARS-CoV-2 Cell Entry Depends on ACE2 and TMPRSS2 and Is Blocked by a Clinically Proven Protease Inhibitor. Cell 2020; 181(2): 271-80.e8.

7. Sungnak W, Huang N, Bécavin C, et al. SARS-CoV-2 entry factors are highly expressed in nasal epithelial cells together with innate immune genes. Nature medicine 2020; 26(5): 681–7.

8. Muus C, Luecken MD, Eraslan G, et al. Integrated analyses of single-cell atlases reveal age, gender, and smoking status associations with cell type-specific expression of mediators of SARS-CoV-2 viral entry and highlights inflammatory programs in putative target cells. bioRxiv 2020: 2020.04.19.049254.

9. Ziegler CGK, Allon SJ, Nyquist SK, et al. SARS-CoV-2 Receptor ACE2 Is an Interferon-Stimulated Gene in Human Airway Epithelial Cells and Is Detected in Specific Cell Subsets across Tissues. Cell 2020; 181(5): 1016-35.e19.

10. Li C, Ji F, Wang L, et al. Asymptomatic and Human-to-Human Transmission of SARS-CoV-2 in a 2-Family Cluster, Xuzhou, China. Emerging infectious diseases 2020; 26(7).

11. Rothe C, Schunk M, Sothmann P, et al. Transmission of 2019-nCoV Infection from an Asymptomatic Contact in Germany. The New England journal of medicine 2020; 382(10): 970–1.

12. Bradley BT, Maioli H, Johnston R, et al. Histopathology and Ultrastructural Findings of Fatal COVID-19 Infections. medRxiv 2020: 2020.04.17.20058545.

13. Wichmann D, Sperhake JP, Lütgehetmann M, et al. Autopsy Findings and Venous Thromboembolism in Patients With COVID-19. Annals of internal medicine 2020.

14. Horton R. Offline: A global health crisis? No, something far worse. Lancet (London, England) 2020; 395(10234): 1410.

15. university JH. Global death from COVID-19. 2020. https://coronavirus.jhu.edu/map.html (accessed 05 july 2020).

16. Guo T, Fan Y, Chen M, et al. Cardiovascular Implications of Fatal Outcomes of Patients With Coronavirus Disease 2019 (COVID-19). JAMA cardiology 2020.

17. Zhou F, Yu T, Du R, et al. Clinical course and risk factors for mortality of adult inpatients with COVID-19 in Wuhan, China: a retrospective cohort study. Lancet (London, England) 2020; 395(10229): 1054–62.

18. Liu Y, Yan LM, Wan L, et al. Viral dynamics in mild and severe cases of COVID-19. The Lancet Infectious diseases 2020; 20(6): 656–7.

19. Thevarajan I, Nguyen THO, Koutsakos M, et al. Breadth of concomitant immune responses prior to patient recovery: a case report of non-severe COVID-19. Nature medicine 2020; 26(4): 453–5.

20. Zhou Z, Ren L, Zhang L, et al. Heightened Innate Immune Responses in the Respiratory Tract of COVID-19 Patients. Cell host & microbe 2020.

21. Huang C, Wang Y, Li X, et al. Clinical features of patients infected with 2019 novel coronavirus in Wuhan, China. Lancet (London, England) 2020; 395(10223): 497–506.

22. Chen G, Wu D, Guo W, et al. Clinical and immunological features of severe and moderate coronavirus disease 2019. The Journal of clinical investigation 2020; 130(5): 2620–9.

23. Liu B, Han J, Cheng X, et al. Persistent SARS-CoV-2 presence is companied with defects in adaptive immune system in non-severe COVID-19 patients. medRxiv 2020: 2020.03.26.20044768.

24. Zheng M, Gao Y, Wang G, et al. Functional exhaustion of antiviral lymphocytes in COVID-19 patients. Cellular & molecular immunology 2020; 17(5): 533–5.

25. Tan L, Wang Q, Zhang D, et al. Lymphopenia predicts disease severity of COVID-19: a descriptive and predictive study. Signal transduction and targeted therapy 2020; 5(1): 33.

26. Grillet F, Behr J, Calame P, Aubry S, Delabrousse E. Acute Pulmonary Embolism Associated with COVID-19 Pneumonia Detected by Pulmonary CT Angiography. Radiology 2020: 201544.

27. Helms J, Tacquard C, Severac F, et al. High risk of thrombosis in patients with severe SARS-CoV-2 infection: a multicenter prospective cohort study. Intensive care medicine 2020: 1–10.

28. Klok FA, Kruip M, van der Meer NJM, et al. Incidence of thrombotic complications in critically ill ICU patients with COVID-19. Thrombosis research 2020; 191: 145–7.

29. Poor HD, Ventetuolo CE, Tolbert T, et al. COVID-19 critical illness pathophysiology driven by diffuse pulmonary thrombi and pulmonary endothelial dysfunction responsive to thrombolysis. Clinical and Translational Medicine; n/a(n/a).

30. Whyte CS, Morrow GB, Mitchell JL, Chowdary P, Mutch NJ. Fibrinolytic abnormalities in acute respiratory distress syndrome (ARDS) and versatility of thrombolytic drugs to treat COVID-19. Journal of thrombosis and haemostasis : JTH 2020.

31. Bhayana R, Som A, Li MD, et al. Abdominal Imaging Findings in COVID-19: Preliminary Observations. Radiology 2020: 201908.

32. Oxley TJ, Mocco J, Majidi S, et al. Large-Vessel Stroke as a Presenting Feature of Covid-19 in the Young. The New England journal of medicine 2020; 382(20): e60.

33. Tang N, Bai H, Chen X, Gong J, Li D, Sun Z. Anticoagulant treatment is associated with decreased mortality in severe coronavirus disease 2019 patients with coagulopathy. Journal of thrombosis and haemostasis : JTH 2020; 18(5): 1094–9.

34. Lakdawala SS, Menachery VD. The search for a COVID-19 animal model. Science (New York, NY) 2020; 368(6494): 942–3.

35. Moher D, Liberati A, Tetzlaff J, Altman DG. Preferred reporting items for systematic reviews and meta-analyses: the PRISMA statement. PLoS medicine 2009; 6(7): e1000097.

36. Munster VJ, Feldmann F, Williamson BN, et al. Respiratory disease in rhesus macaques inoculated with SARS-CoV-2. Nature 2020.

37. Yu P, Qi F, Xu Y, et al. Age-related rhesus macaque models of COVID-19. Animal models and experimental medicine 2020; 3(1): 93–7.

38. Yu J, Tostanoski LH, Peter L, et al. DNA vaccine protection against SARS-CoV-2 in rhesus macaques. Science (New York, NY) 2020.

39. van Doremalen N, Lambe T, Spencer A, et al. ChAdOx1 nCoV-19 vaccination prevents SARS-CoV-2 pneumonia in rhesus macaques. bioRxiv 2020: 2020.05.13.093195.

40. Gao Q, Bao L, Mao H, et al. Development of an inactivated vaccine candidate for SARS-CoV-2. Science (New York, NY) 2020: eabc1932.

41. Williamson BN, Feldmann F, Schwarz B, et al. Clinical benefit of remdesivir in rhesus macaques infected with SARS-CoV-2. bioRxiv 2020: 2020.04.15.043166.

42. Chandrashekar A, Liu J, Martinot AJ, et al. SARS-CoV-2 infection protects against rechallenge in rhesus macaques. Science (New York, NY) 2020.

43. Bao L, Deng W, Gao H, et al. Lack of Reinfection in Rhesus Macaques Infected with SARS-CoV-2. bioRxiv 2020: 2020.03.13.990226.

44. Deng W, Bao L, Gao H, et al. Ocular conjunctival inoculation of SARS-CoV-2 can cause mild COVID-19 in Rhesus macaques. bioRxiv 2020: 2020.03.13.990036.

45. Lu S, Zhao Y, Yu W, et al. Comparison of SARS-CoV-2 infections among 3 species of non-human primates. bioRxiv 2020: 2020.04.08.031807.

46. Finch CL, Crozier I, Lee JH, et al. Characteristic and quantifiable COVID-19-like abnormalities in CT- and PET/CT-imaged lungs of SARS-CoV-2-infected crab-eating macaques (<em>Macaca fascicularis</em>). bioRxiv 2020: 2020.05.14.096727.

47. Rockx B, Kuiken T, Herfst S, et al. Comparative pathogenesis of COVID-19, MERS, and SARS in a nonhuman primate model. Science (New York, NY) 2020; 368(6494): 1012–5.

48. Woolsey C, Borisevich V, Prasad AN, et al. Establishment of an African green monkey model for COVID-19. bioRxiv 2020: 2020.05.17.100289.

49. Bao L, Deng W, Huang B, et al. The pathogenicity of SARS-CoV-2 in hACE2 transgenic mice. Nature 2020.

50. Li W, Drelich A, Martinez DR, et al. Rapid selection of a human monoclonal antibody that potently neutralizes SARS-CoV-2 in two animal models. bioRxiv 2020: 2020.05.13.093088.

51. Dinnon KH, Leist SR, Schäfer A, et al. A mouse-adapted SARS-CoV-2 model for the evaluation of COVID-19 medical countermeasures. bioRxiv 2020: 2020.05.06.081497.

52. Lv H, Wu NC, Tsang OT, et al. Cross-reactive Antibody Response between SARS-CoV-2 and SARS-CoV Infections. Cell reports 2020; 31(9): 107725.

53. Gu H, Chen Q, Yang G, et al. Rapid adaptation of SARS-CoV-2 in BALB/c mice: Novel mouse model for vaccine efficacy. bioRxiv 2020: 2020.05.02.073411.

54. Pruijssers AJ, George AS, Schäfer A, et al. Remdesivir potently inhibits SARS-CoV-2 in human lung cells and chimeric SARS-CoV expressing the SARS-CoV-2 RNA polymerase in mice. bioRxiv 2020: 2020.04.27.064279.

55. Boudewijns R, Thibaut HJ, Kaptein SJF, et al. STAT2 signaling as double-edged sword restricting viral dissemination but driving severe pneumonia in SARS-CoV-2 infected hamsters. bioRxiv 2020: 2020.04.23.056838.

56. Chan JF-W, Zhang AJ, Yuan S, et al. Simulation of the clinical and pathological manifestations of Coronavirus Disease 2019 (COVID-19) in golden Syrian hamster model: implications for disease pathogenesis and transmissibility. Clinical Infectious Diseases 2020.

57. Rogers TF, Zhao F, Huang D, et al. Rapid isolation of potent SARS-CoV-2 neutralizing antibodies and protection in a small animal model. bioRxiv 2020: 2020.05.11.088674.

58. Sia SF, Yan LM, Chin AWH, et al. Pathogenesis and transmission of SARS-CoV-2 in golden hamsters. Nature 2020.

59. Kim YI, Kim SG, Kim SM, et al. Infection and Rapid Transmission of SARS-CoV-2 in Ferrets. Cell host & microbe 2020; 27(5): 704-9.e2.

60. Blanco-Melo D, Nilsson-Payant BE, Liu WC, et al. Imbalanced Host Response to SARS-CoV-2 Drives Development of COVID-19. Cell 2020; 181(5): 1036-45.e9.

61. Richard M, Kok A, de Meulder D, et al. SARS-CoV-2 is transmitted via contact and via the air between ferrets. bioRxiv 2020: 2020.04.16.044503.

62. Shi J, Wen Z, Zhong G, et al. Susceptibility of ferrets, cats, dogs, and other domesticated animals to SARS-coronavirus 2. Science (New York, NY) 2020; 368(6494): 1016–20.

63. Zhao Y, Wang J, Kuang D, et al. Susceptibility of tree shrew to SARS-CoV-2 infection. bioRxiv 2020: 2020.04.30.029736.

64. Hanley B, Lucas SB, Youd E, Swift B, Osborn M. Autopsy in suspected COVID-19 cases. Journal of Clinical Pathology 2020; 73(5): 239–42.

65. Grasselli G, Zangrillo A, Zanella A, et al. Baseline Characteristics and Outcomes of 1591 Patients Infected With SARS-CoV-2 Admitted to ICUs of the Lombardy Region, Italy. Jama 2020; 323(16): 1574–81.

66. Subbarao K, Roberts A. Is there an ideal animal model for SARS? Trends in microbiology 2006; 14(7): 299–303.

67. Sutton TC, Subbarao K. Development of animal models against emerging coronaviruses: From SARS to MERS coronavirus. Virology 2015; 479-480: 247–58.

68. Ackermann M, Verleden SE, Kuehnel M, et al. Pulmonary Vascular Endothelialitis, Thrombosis, and Angiogenesis in Covid-19. The New England journal of medicine 2020.

69. Marini JJ, Gattinoni L. Management of COVID-19 Respiratory Distress. Jama 2020.

70. Xu Z, Shi L, Wang Y, et al. Pathological findings of COVID-19 associated with acute respiratory distress syndrome. The Lancet Respiratory medicine 2020; 8(4): 420–2.

71. Nicholls JM, Poon LL, Lee KC, et al. Lung pathology of fatal severe acute respiratory syndrome. Lancet (London, England) 2003; 361(9371): 1773–8.

72. Varga Z, Flammer AJ, Steiger P, et al. Endothelial cell infection and endotheliitis in COVID-19. Lancet (London, England) 2020; 395(10234): 1417–8.

73. Mulvey JJ, Magro CM, Ma LX, Nuovo GJ, Baergen RN. Analysis of complement deposition and viral RNA in placentas of COVID-19 patients. Annals of diagnostic pathology 2020; 46: 151530.

74. Gupta N, Zhao YY, Evans CE. The stimulation of thrombosis by hypoxia. Thrombosis research 2019; 181: 77–83.

75. Zhang Y, Xiao M, Zhang S, et al. Coagulopathy and Antiphospholipid Antibodies in Patients with Covid-19. The New England journal of medicine 2020; 382(17): e38.

76. Bowles L, Platton S, Yartey N, et al. Lupus Anticoagulant and Abnormal Coagulation Tests in Patients with Covid-19. The New England journal of medicine 2020.

77. Lee N, Hui D, Wu A, et al. A major outbreak of severe acute respiratory syndrome in Hong Kong. The New England journal of medicine 2003; 348(20): 1986–94.

78. Zaki AM, van Boheemen S, Bestebroer TM, Osterhaus ADME, Fouchier RAM. Isolation of a Novel Coronavirus from a Man with Pneumonia in Saudi Arabia. New England Journal of Medicine 2012; 367(19): 1814–20.

79. Damas J, Hughes GM, Keough KC, et al. Broad Host Range of SARS-CoV-2 Predicted by Comparative and Structural Analysis of ACE2 in Vertebrates. bioRxiv 2020: 2020.04.16.045302.

80. Han L, Wei X, Liu C, et al. Single-cell atlas of a non-human primate reveals new pathogenic mechanisms of COVID-19. bioRxiv 2020: 2020.04.10.022103.

81. Hagai T, Chen X, Miragaia RJ, et al. Gene expression variability across cells and species shapes innate immunity. Nature 2018; 563(7730): 197–202.

